# Imputation of PaO2 from SpO2 values from the MIMIC-III Critical Care Database Using Machine-Learning Based Algorithms

**DOI:** 10.1101/2021.04.21.21255877

**Authors:** Shuangxia Ren, Jill Zupetic, Mehdi Nouraie, Xinghua Lu, Richard D. Boyce, Janet S. Lee

## Abstract

**Background:** The partial pressure of oxygen (PaO2)/fraction of oxygen delivered (FIO2) ratio is the reference standard for assessment of hypoxemia in mechanically ventilated patients. Non-invasive monitoring with the peripheral saturation of oxygen (SpO2) is increasingly utilized to estimate PaO2 because it does not require invasive sampling. Several equations have been reported to impute PaO2/FIO2 from SpO2 /FIO2. However, machine-learning algorithms to impute the PaO2 from the SpO2 has not been compared to published equations.

**Research Question:** How do machine learning algorithms perform at predicting the PaO2 from SpO2 compared to previously published equations?

**Methods:** Three machine learning algorithms (neural network, regression, and kernel-based methods) were developed using 7 clinical variable features (n=9,900 ICU events) and subsequently 3 features (n=20,198 ICU events) as input into the models from data available in mechanically ventilated patients from the Medical Information Mart for Intensive Care (MIMIC) III database. As a regression task, the machine learning models were used to impute PaO2 values. As a classification task, the models were used to predict patients with moderate-to-severe hypoxemic respiratory failure based on a clinically relevant cut-off of PaO2/FIO2 ≤ 150. The accuracy of the machine learning models was compared to published log-linear and non-linear equations. An online imputation calculator was created.

**Results:** Compared to seven features, three features (SpO2, FiO2 and PEEP) were sufficient to impute PaO2/FIO2 ratio using a large dataset. Any of the tested machine learning models enabled imputation of PaO2/FIO2 from the SpO2/FIO2 with lower error and had greater accuracy in predicting PaO2/FIO2 ≤ 150 compared to published equations. Using three features, the machine learning models showed superior performance in imputing PaO2 across the entire span of SpO2 values, including those ≥ 97%.

**Interpretation:** The improved performance shown for the machine learning algorithms suggests a promising framework for future use in large datasets.

## Introduction

The PaO2 as a ratio of the fraction of oxygen (FIO2) delivered, or the PaO2/ FIO2, is the reference standard measurement for assessment of hypoxemia in mechanically ventilated patients with respiratory failure. The PaO2/FIO2 ratio (PF ratio) has predictive value for mortality in patients with acute respiratory distress syndrome (ARDS)^1^ and is also part of a severity index scoring system called the Sequential Organ Failure Assessment (SOFA) score that is used to predict mortality in patients with critical illness^2–4^. Additionally, the PaO2/FIO2 ratio has become relevant in clinical decision-making including the decision to initiate prone positioning in ARDS patients with PF ratios less than 150^5^. Current measurement of the PaO2/FiO2 ratio requires invasive arterial blood gas sampling and does not provide a continuous measure of the patient’s oxygenation. Increasingly, non-invasive monitoring with pulse oximetry is utilized instead of ABGs^6,7^, particularly in low-resource settings where an ABG lab or invasive arterial blood monitoring are not readily available or required. In addition, several studies have evaluated the non-invasive SpO2 (peripheral saturation of oxygen)/FIO2 ratio as a surrogate for PaO2/FIO2 ratio in children where non-invasive measurements are becoming more common^8–10^.

A few studies have examined non-linear imputation of PaO2/FIO2 from SpO2/FIO2 measurements recorded at the same time^11,12^. These studies have reported that the accuracy of non-linear imputation is superior to log-linear or linear imputation, especially for moderate to severe hypoxemic respiratory failure with ARDS^11,13^. However, in patients with respiratory failure requiring mechanical ventilation, the optimal equation for imputation of PaO2/FIO2 from the SpO2/FIO2 remains unclear. An algorithm to accurately impute the PaO2 from the SpO2 in mechanically ventilated patients would be beneficial for clinical research to facilitate recruitment of patients for clinical trials if an ABG is not available. Ideally, this approach would involve only the introduction of variables that may contribute to the relationship between SpO2 and PaO2 but would not require the same invasive ABG measurement as the PaO2.

The objective of this study is to develop a machine learning algorithm to impute PaO2/FIO2 from SpO2/FIO2 among mechanically ventilated patients in the Medical Information Mart for Intensive Care (MIMIC) III database^14^ and compare it to the previously published non-linear and log-linear equations^11,13^. In this study, three common machine learning approaches (neural network^15^, regression^16^, and kernel-based methods^17,18^) were tested for regression and classification of PaO2/FIO2 using data available in MIMIC III^19^ with 7 clinical variable features and a subsequent 3 features model. We created models to perform a regression task to impute PaO2 from SpO2 values and a classification task to predict patients with moderate to severe hypoxemic respiratory failure based on a cut-off of a predicted PF ratio ≤ 150^11^. Our overall hypothesis was that a machine learning algorithm would perform better in predicting the PaO2 from SpO2 across the entire span of SpO2 values when compared to the published equations.

## Methods

The MIMIC-III database v1.4 (https://mimic.physionet.org) is an openly available dataset developed by the Massachusetts Institute of Technology Lab for Computational Physiology^14^. It contains de-identified health data associated with approximately 60,000 intensive care unit admissions. MIMIC-III is a relational database that contains information on demographics, vital signs, mechanical ventilation status, laboratory tests, medications, and mortality. Our study was determined by the University of Pittsburgh Institutional Review Board to be exempt (STUDY19100068).

### Data processing

We queried the MIMIC-III database to identify unique ICU encounters (icustay_id) with mechanical ventilation status. We next identified the lab event PaO2 and chart event SpO2 occurring at the same time of the mechanical ventilation status. In order to minimize error between matched PaO2 and SpO2, we constrained the time gap between the lab event PaO2 and the chart event SpO2 to the closest time within 30 minutes. To minimize repeated sampling from the same subjects, we restricted the search of PaO2 measurements to within the first 24 hours of mechanical ventilation duration and obtained the first PaO2 recorded within this time frame. We constrained the time gap to within 2 hours of the selected SpO2 measurement for variables from chart events such as tidal volume (TV), positive end-expiratory pressure (PEEP), FiO2, temperature, and mean arterial pressure (MAP). We did the same for lab events such as SaO2. If a patient was treated with vasoactive infusions, it was recorded as a categorical variable. Data extraction and processing methods are available at https://github.com/renshuangxia/Predict-PaO2-with-SpO2^20^.

### Machine learning methods for regression task

For the regression task we implemented 3 different models – a neural network model, a linear regression model, and support vector regression (SVR), a type of kernel-based modeling. For each model, we applied a 10-fold cross-validation^21^.

For the neural network model, we tested different network structures and various numbers of features to arrive at two models used for comparison with the linear and support vector regression models. One model used seven input features and three hidden layers (16, 8, 5 neurons for layers 1 to 3). The other model used only three input features and two hidden layers (6, 3 neurons for layers 1 and 2). Both final models used a tangent activation function for all layers except the output layer which used a linear function in both models. Also, both models were trained for 200 epochs with Adam optimizer using gradient descent. The learning rate was 0.001 and the batch sizes were 50 for both models.

For the linear regression model, the output variable can be computed by a linear combination of the input variables. We trained the linear regression equation by the Ordinary Least Squares approach. We used the linear_model.LinearRegression method from scikit-learn 0.22 (https://scikit-learn.org/stable/) with default hyperparameters for predicting PaO2 values.

For the SVR model, we tested multiple kernels including linear kernel, polynomial kernel, and radical basis function kernel (RBF). Based on the performance in the training data, the RBF kernel was selected.

### Machine learning methods for classification task

In patients meeting criteria for ARDS, the PaO2/FIO2 ≤ 150 has been used to capture those patients with moderate to severe disease^11,13^. We utilized this cut-off to test machine learning methods to predict this diagnostic threshold PaO2/FIO2 ≤ 150 for the different imputation techniques. We implemented 3 classification models including Neural Network, Logistic Regression and Support Vector Machine (SVM).

For each of machine learning model we applied a 10-fold cross-validation and calculated the sensitivity, specificity, likelihood ratios, diagnostic odds ratio (OR), Area under receiver operating characteristic curve (AUROC), F1 score and Bayesian information Criterion (BIC) to compare across models. The two neural network models for classification were similar to the neural networks used in regression, except the output layer used the sigmoid function. As with the regression models, various topologies were tested to arrive at the final two multi-layer perceptron (MLP) classifiers, one with an input size of 7 features and the other with an input size of 3 features. The hidden layer size is (12, 8, 6, 4, 4) for the model with 7 input features. For the other model which utilizes only 3 input features, we used two hidden layers of size 6 and 3. All hidden layers used the tangent activation function. We trained both models for 200 iterations with Adam optimizer, setting 7 feature classifier momentum value as 0.8 and 3 feature classifier momentum value as 0.6. The learning rate was 0.001 and the batch sizes was 200 for both models.

In addition, we implemented a basic logistic regression model for classification purposes as well as the SVM model which classifies examples with an optimal hyperplane. For the logistic regression, it uses logistic function to model a binary dependent variable. We utilized the linear_model.LogisticRegression method provided in the scikit-learn library without regularization, and other arguments were set as default. For the SVM model, we compared the results by applying different kernels and the RBF kernel outperformed other kernels. Methods were similar to those used in the regression task.

### Comparison of machine-learning based algorithm to published non-linear and log-linear equations

We compared the performance of our machine learning algorithms to the previously published equations. For the non-linear equation from Brown *et al*^11^ the PaO2 was imputed from the SpO2, where PO_2_ = PaO_2_, S = SpO_2_ and F=FiO2. For situations where the recorded SpO2 was 100% (or, 1.0), the SpO2 was substituted with 0.996 given that the equation would not permit the calculation of S=1.0.

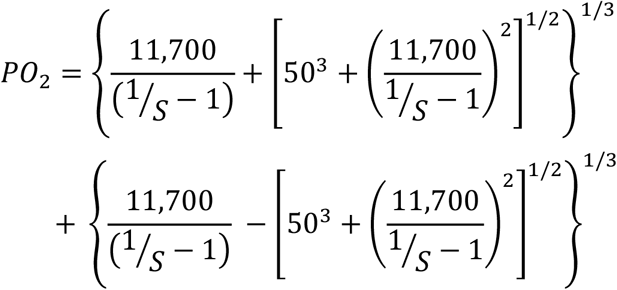

For the log-linear equation from Pandharipande, et al^11,13^, the PaO_2_:FIO2 was imputed from SpO_2_:FIO2 utilizing the equation:

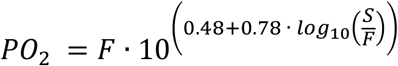

## Results

### A parsimonious three features model is sufficient to impute PaO2/FIO2 ratio using a large dataset

An overview of the machine learning tasks are outlined in Figure 1. We initially chose 7 relevant features from the chart events (SpO2, FiO2, TV, MAP, temperature, PEEP and vasopressor administration) representing recorded bedside measurements that were independent from an invasive arterial blood gas measurement. When applying the 7 features to impute PaO2/FiO2, the final data set contained 9,900 unique ICU encounters from 9,302 mechanically ventilated patients (Supplementary Table e1). The relationship between SpO2/FiO2 (S/F) and the PaO2/FiO2 (P/F) was examined in dataset 1 containing 9,900 unique ICU events from the MIMIC-III database and was best described by a log-linear relationship between the transformed logarithmic value of the SF and PF ratios as previously described by Pandharipande, et al^13^ (Supplementary Figure e1). The relationship between S/F and P/F ratios showed high variance across the distribution of mechanically ventilated subjects (R^2^ = 0.21).

**Figure 1.**
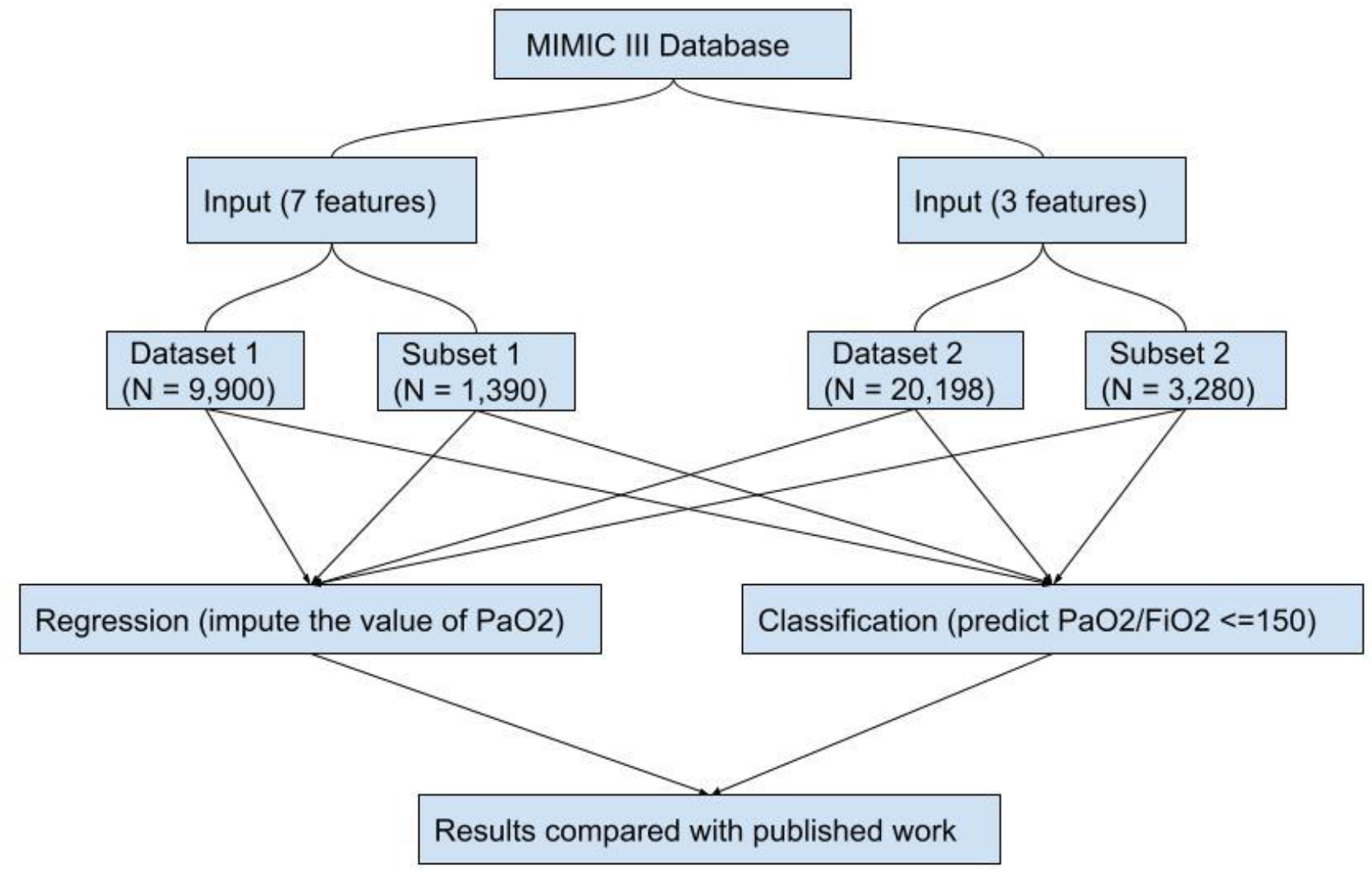
Overview of the experimental study design.

For the regression task, we derived the RMSE and BIC for each of the different 7 feature machine learning models (neural network, linear regression, support vector regression) to assess the performance of the imputation techniques. The RMSE and BIC of the three machine learning methods are shown in Supplementary Table e2. All the machine learning models outperformed the previously published non-linear and log-linear equations as shown by lower RMSE scores. For the classification task, the three machine learning methods achieved similar classification performance according to F1 scores, as shown in Supplementary Table e3.

To improve practicality of the method at the bedside, we attempted to use the smallest number of features possible to predict PaO2 or PaO2/FiO2 ratio from the regression and classification tasks, respectively. Compared to the other measured variables, PEEP had the strongest correlation with PaO2/FiO2 (r = −0.31) outside of the SF ratio (SpO2/FiO2) (Table 1). Using this information, we created a 3-features model using SpO2, FIO2 and PEEP. As compared to seven features, three features were sufficient to impute PaO2/FIO2 ratio with a similar degree of accuracy in part due to the ability to include significantly more subjects. The 3 features model was therefore utilized in the remainder of the analysis. The final 3 features data set (dataset 2) contained 20,198 ICU encounters from 17,818 unique patients (Table 2). Forty percent of subjects were of female sex and the mean age was 64 years. The degree of hypoxemic respiratory failure, as measured by the PaO2/FIO2 ratio^1^, showed a distribution in which 26% had mild respiratory failure (PaO2/FIO2 = 201-300), 22% had moderate respiratory failure (PaO2/FIO2 = 101-200), and 8% had severe respiratory failure (PaO2/FIO2 < 100).

**Table 1.**
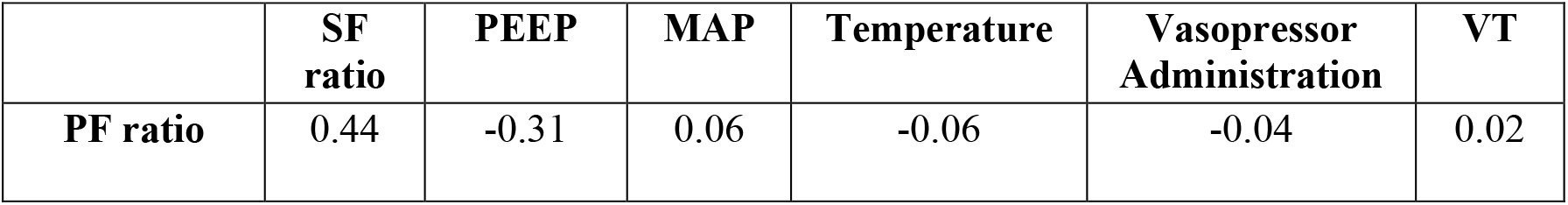
Correlation coefficients between PF ratios and variables. Correlation coefficients between measured PF ratios (PaO2/FiO2) and the 6 other measured variables (SpO2/FiO2 = SF ratio, PEEP, MAP, Temperature, Vasopressor Administration and VT) were performed. The variable with the strongest correlation coefficient (r) was chosen for the 3 features model.

**Table 2.** Subject characteristics based on 3 features. The 3 features models captured 20,198 ICU events from 17,818 unique patients. Variables included in the 3 features machine learning models are SpO2, FiO2, and PEEP. ^*^For subjects older than 89 years, the age was assigned as 90 years of age.

|  |  |
| --- | --- |
| Total ICU events, N | 20,198 |
| Female sex, n (%) | 8,084 (40.0) |
| Age in years, mean (± SD)* | 64.0 (± 16.2) |
| PaO<sub>2</sub>/FIO<sub>2</sub>, mean (± SD) | 310.4 (±184.4) |
| Available mean PaO<sub>2</sub>/FIO<sub>2</sub>, N | 20,198 |
| PaO<sub>2</sub>/FIO<sub>2</sub> >300, n | 8996 |
| PaO<sub>2</sub>/FIO<sub>2</sub> = 201-300, n | 5226 |
| PaO<sub>2</sub>/FIO<sub>2</sub> = 101-200, n | 4448 |
| PaO<sub>2</sub>/FIO<sub>2</sub> < 100, n | 1528 |
| Available SpO<sub>2</sub> measurements per unique patient, N | 17,818 |
| 1 measurement, n | 16,065 |
| 2 measurements, n | 1,367 |
| 3 measurements, n | 262 |
| 4 measurements, n | 77 |
| 5 measurements, n | 29 |
| 6 measurements, n | 14 |
| 7 measurements, n | 4 |

Abbreviations: PEEP= Positive end- expiratory pressure.

### Machine learning models show improved performance when compared to the prior published equations for regression

We quantitatively derived the RMSE for all the machine learning and previously published models and the BIC for each of the three machine learning models to assess the performance of the different imputation techniques (Table 3). The RMSE of the neural network, linear regression and support vector regression machine learning models were 84.7, 88.8 and 85.9, respectively, compared to 117.7 and 91.8 for the log-linear and non-linear equations. The lower RMSE values indicate that the 3 machine learning models outperformed the previously published equations. Of the machine learning models, the neural network method showed the lowest RMSE as well as the lowest BIC in both the whole dataset (dataset 2) and for Sp02 <97% (subset 2), and thus was chosen as the “best” overall model for the regression task. A Bland-Altman Plot suggests that the neural network model is comparable to the published equations. There was decreasing accuracy at higher PaO2/FIO2 ratios for all the methods examined (Supplementary Figure e2).

**Table 3.**
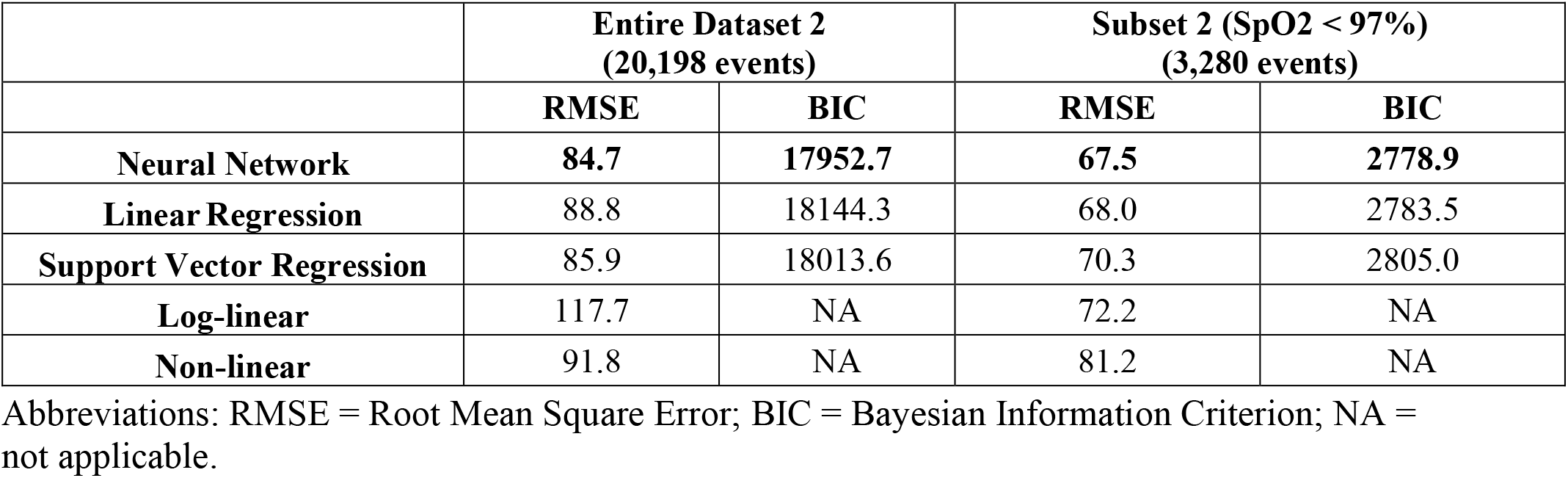
RMSE and BIC of the 3 features machine learning models regression tasks compared to published methods. The RMSE and BIC for the 3 features machine learning models were calculated for the entire dataset (20,198 ICU events) and a subset of the dataset with SpO2 < 97% (3,280 ICU events) and compared to the published log-linear and non-linear models.

|  | **Entire Dataset 2**<br>**(20,198 events)** |  | **Subset 2 (SpO2 < 97%)**<br>**(3,280 events)** |  |
| --- | --- | --- | --- | --- |
|  | **RMSE** | **BIC** | **RMSE** | **BIC** |
| **Neural Network** | **84.7** | **17952.7** | **67.5** | **2778.9** |
| **Linear Regression** | 88.8 | 18144.3 | 68.0 | 2783.5 |
| **Support Vector Regression** | 85.9 | 18013.6 | 70.3 | 2805.0 |
| **Log-linear** | 117.7 | NA | 72.2 | NA |
| **Non-linear** | 91.8 | NA | 81.2 | NA |

Abbreviations: RMSE = Root Mean Square Error; BIC = Bayesian Information Criterion; NA = not applicable.

### Machine learning models show improved performance for the classification task

We compared the performance of the machine learning models with the log-linear and non-linear equations using F1 scores. Similar to the findings for the regression task, all three machine learning models performed better in the whole dataset than log-linear and non-linear equations (Table 4). When the dataset was limited to SpO2 < 97% (subset 2), the machine-learning methods performed slightly better than log-linear and better than non-linear equations, respectively (Table 4). The F1 scores for all three machine learning methods were similar when using the whole dataset (dataset 2) and for subset 2 where SpO2 < 97%. As shown in Figure 2, when comparing the 3 machine learning models to one another, the neural network preformed slightly better in the whole dataset (area under the precision recall curve = 0.94 for the neural network compared to 0.93 and 0.91 for the logistic regression and support vector machine model, respectively). The 3 models had similar performance in subset 2.

**Table 4.** Prediction performance of machine learning classification models based on 3 features. Prediction performance statistics were calculated for the machine learning models based on 3 features and compared to the Log-linear and Non-linear methods for the entire dataset (20,198 ICU events; entire dataset 2) and for a subset of the events where SpO2 <97% (3,280 events; subset 2). Variables included in the 3 features machine learning models are SpO2, FiO2, and PEEP.

|  | Entire Dataset 2<br>(20,198 events) |  |  |  |  | Subset 2 (SpO2 < 97%)<br>(3,280 events) |  |  |  |  |
| --- | --- | --- | --- | --- | --- | --- | --- | --- | --- | --- |
|  | Neural<br>Network | Logistic<br>Regression | SVM | Log-linear | Non-linear | Neural<br>Network | Logistic<br>regression | SVM | Log-linear | Non-linear |
| **Total, No.** | 20198 | 20198 | 20198 | 20198 | 20198 | 3280 | 3280 | 3280 | 3280 | 3280 |
| **Sensitivity** | 0.96 | 0.97 | 0.98 | 0.84 | 0.93 | 0.80 | 0.87 | 0.83 | 0.85 | 0.58 |
| **Specificity** | 0.39 | 0.26 | 0.33 | 0.56 | 0.49 | 0.76 | 0.59 | 0.69 | 0.59 | 0.89 |
| **Positive**<br>**LR** | 1.59 | 1.32 | 1.46 | 1.90 | 1.83 | 3.37 | 2.13 | 2.75 | 2.09 | 5.16 |
| **Negative**<br>**LR** | 0.09 | 0.10 | 0.07 | 0.29 | 0.15 | 0.27 | 0.23 | 0.25 | 0.25 | 0.47 |
| **Diagnostic**<br>**OR** | 17.12 | 13.16 | 19.68 | 6.49 | 12.61 | 12.53 | 9.46 | 10.96 | 8.44 | 10.94 |
| **AUROC** | **0.83** | 0.81 | 0.74 | NA | NA | **0.85** | 0.83 | 0.84 | NA | NA |
| **F1** | 0.92 | 0.92 | 0.92 | 0.87 | 0.91 | **0.81** | 0.80 | 0.81 | 0.79 | 0.70 |
| **BIC** | **-4612.60** | -4440.70 | -4446.00 | NA | NA | **-591.80** | -567.00 | -580.00 | NA | NA |

Abbreviations: SVM = Support Vector Machine; Positive LR = Positive Likelihood Ratio; Negative LR = Negative Likelihood Ratio; Diagnostic OR = Diagnostic Odds Ratio (Ratio of Positive Likelihood Ratio/ Negative Likelihood Ratio); AUROC = Area Under Receiver Operating Characteristic Curve; F1= F1 score; BIC = Bayesian Information Criterion; NA = Not applicable.

**Figure 2:**
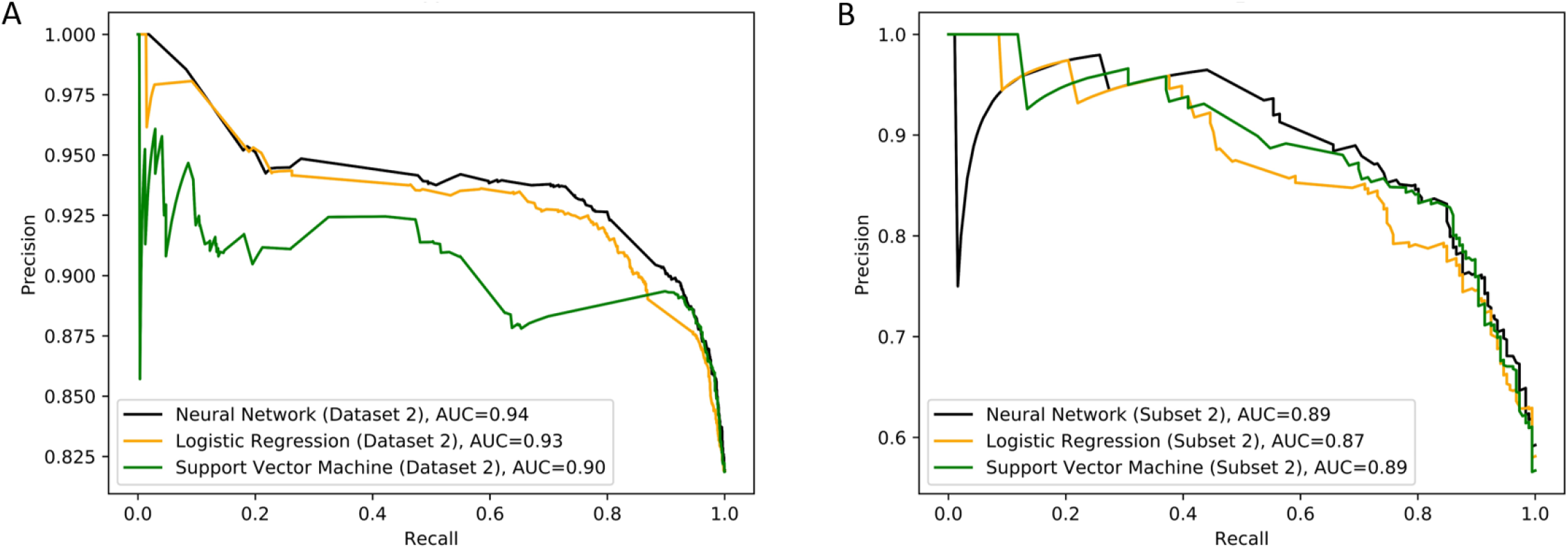
Precision-recall curves of machine learning models in Dataset 2 and Subset 2 using 3 features. The precision recall curves, where improved performance is demonstrated if the curve is closer to the upper right-hand corner or has the highest area under the curve (AUC), are shown for the 3 machine learning models for A) the entire Dataset 2 (N = 20,198) ICU events) and B) Subset 2 where SpO2 <97% (N = 3,280 ICU events).

## Discussion

We used the publicly available MIMIC-III database to develop and evaluate machine-learning algorithms to impute PaO2/FIO2 from the SpO2/FIO2 in patients who are mechanically ventilated. We tested three machine learning models (neural network, linear regression and SVR) first using seven available clinical variables SpO2, FIO2, PEEP, TV, MAP, temperature, and vasopressor administration to impute the PaO2 and subsequently using only three clinical variables SpO2, FiO2 and PEEP. The imputation of PaO2 from the SpO2 from the regression tasks enabled us to derive the PaO2/FIO2, a clinically meaningful ratio with predictive value^1,22^. Additionally, we performed a classification task to predict PaO2/FiO2 ≤ 150, a cut off that has been used to capture those patients with moderate to severe respiratory failure in ARDS cohorts^11,13^ and to guide patient management^5^.

To develop the machine learning algorithms, we evaluated clinical variables such as PEEP, TV, MAP, temperature, and vasopressor administration that are easily obtained at the bedside. We considered other clinical variables such as skin pigmentation, pulse oximeter location, oximeter manufacturer, vasopressor infusion, and laboratory variables such as serum bicarbonate, serum chloride, serum creatinine, serum sodium but these variables added negligible improvement in the accuracy of imputation in a prospective study^11^. Therefore, these additional clinical variables were not added to the model. Except for PEEP, other variables examined showed a stochastic distribution. Removing these unrelated features (VT, MAP, Temperature and vasopressor use) to create the 3 features model did not significantly alter the accuracy of the machine-learning based algorithms and provides a framework for the generalizability of the model for large datasets of mechanically ventilated patients.

Our study shows that a machine-learning based method for both the regression and classification task, when applied to the MIMIC-III critical care database, improved the accuracy when compared with the prior published non-linear, and log-linear imputation methods. As is evidenced by comparing the F1 and discrimination measures in Table 4, the performance improvement was more modest for the classification task in subset 2 where SpO2 <97%. A possible explanation is that there were fewer ICU events (smaller N) per group in the subset.

Prior studies have examined the relationship between SpO2/FIO2 (SF) and PaO2/FIO2 (PF) ratios for patients with ARDS to determine whether the non-invasive SF ratio can be substituted for the invasively obtained PF ratio^11,13,23^. Panharipande, et al studied matched measurements of SpO2 and PaO2 of a more heterogenous population to determine the association between SF and PF ratios in order to calculate the respiratory parameter of the SOFA score^13^. In their study, matched SpO2 and PaO2 values were obtained from two groups of patients: Group 1 comprised of the derivation set and was obtained from patients undergoing general anesthesia from a single center, and Group 2 comprised of a validation set and was obtained from patients enrolled in the multi-center randomized clinical trial examining low versus high tidal volume for acute respiratory management of ARDS (ARMA)^24^. All SpO2 values > 97% were also excluded from analysis in order to maximize matched data to those values likely to be within the linear range of the oxyhemoglobin dissociation curve. Data from 4,728 matched SpO2 and PaO2 measurements showed that the relationship was best described by a log-linear equation with slight variation based upon the level of PEEP. In the setting of a more heterogenous population, a poorer correlation was noted between SF and PF ratios. The regression equation of Log(PF) = 0.48+0.78 x Log(SF) yielded an R-square of 0.31^13^.

A retrospective analysis of enrollment arterial blood gas measurements from three ARDS Network studies compared the performance of non-linear, log-linear and linear imputation methods to derive PaO2 from the SpO2^12^. In all patients (N=1,184), the nonlinear imputation was equivalent to log-linear imputation. However, in those patients with SpO2 < 97% (N=707), the nonlinear imputation showed lower error than either linear or log-linear equations. A prospective study was subsequently conducted in patients enrolled in the Prevention and Early Treatment of Acute Lung Injury network^11^ to assess the performance of the non-linear equation to impute PaO2 from the SpO2 and compare it to the prior log-linear and linear equations^11,13,23^. This study included 1034 arterial blood gases from 703 patients, of which 650 arterial blood gases had matched SpO2 < 97%. The non-linear equation showed lower error and better identified moderate to severe ARDS patients (defined in the study as PaO2/FIO2 ≤ 150) when compared to log-linear or linear imputation methods.

In our study, we similarly found a high degree of variance across SpO2 values and corresponding measured PaO2 values which was noted when we formally examined the relationship between SpO2/FIO2 and PaO2/FIO2. This may be attributed to the retrospective nature of the data collection and the numerous variables that may confound the reliability of a recorded SpO2 measured non-invasively to reflect the arterial SaO2^8,10,12^. Despite this limitation, the machine learning algorithms performed better on both regression and classification tasks when compared to the log-linear and non-linear published equations.

One strength of our study is the evaluation of all mechanically ventilated patients with available data rather than narrowing the analysis to a specific population such as those with ARDS. Given the inclusion of all mechanically ventilated patients, a significant number of SpO2 values were > 97% (N=8,510 for 7 features and N=16,918 for 3 features). While this reduced the accuracy of the imputed PF ratio, particularly above a certain threshold, the machine learning models were applied to the data without a pre-defined restriction placed upon the range of SpO2 values and showed better performance than both the log-linear and non-linear equations on both the regression and classification tasks. These results have not been tested on data other than MIMIC-III. Future work will need to test if the model is robust given potential variations in how the data for input features is collected and stored.

In summary, any of the tested machine learning models applied to MIMIC-III enabled imputation of PaO2/FIO2 ratio from the SpO2/FIO2 with lower error and greater accuracy in predicting PaO2/FIO2 ≤ 150 than when compared to that of published equations across the entire range of SpO2 examined. When compared to one another, all machine learning methods performed similarly. Given our goal of utilizing this type of modeling to allow for inclusion of mechanically ventilated patients from large datasets in the electronic health record, we opted to create a calculator for the neural network machine learning algorithm based on ease of utilization https://drive.google.com/drive/folders/1AoieWO0w3BXvEpw6c0-OomjeQHzFJXY_?usp=sharing. Future studies will need to assess the generalizability of and improve upon the machine learning methods in mechanically ventilated patients to impute PaO2/FIO2 measurements from SpO2 values.

## Data Availability

The data utilized in this manuscript was obtained from the MIMIC-III database v1.4. The MIMIC-III database is an openly available dataset developed by the Massachusetts Institute of Technology Lab for Computational Physiology that contains de-identified health data associated with approximately 60,000 intensive care unit admissions.

https://mimic.physionet.org

## Authorship

### Contribution

S.R. performed the data extraction and processing, analysis, and interpreted the data. J.Z. performed data analysis, interpreted the data and wrote the manuscript. R.B. and X.L. interpreted the data and revised the work for important intellectual content. M.N. provided critical statistical expertise, designed, analyzed, interpreted the data, and wrote the manuscript. J.S.L. conceived, designed, analyzed, interpreted the data, and wrote the manuscript. S.R. and J.Z are the guarantors of the paper.

## Abbreviations List

PaO2: partial pressure of oxygen
FIO2: fraction of oxygen
PaO2/FIO2: PF ratio
SpO2: peripheral saturation of oxygen
PEEP: positive end expiratory pressure
TV: tidal volume
MAP: mean arterial pressure

