## Supplementary Table e1 for "Imputation of PaO2 from SpO2 values from the MIMIC-III Critical Care Database Using Machine-Learning Based Algorithms"

| Total ICU events, N | 9,900 |
| --- | --- |
| Female sex, n (%) | 3,713 (37.5) |
| Age in years, mean (± SD)^*^ | 64.0 (± 16) |
| PaO2/FIO2, mean (± SD) | 322.8 (±193.0) |
| Available mean PaO2/FIO2, N | 9,900 |
| PaO2/FIO2 > 300, n | 4,656 |
| PaO2/FIO2 = 201-300, n | 2,468 |
| PaO2/FIO2 = 101-200, n | 2,042 |
| PaO2/FIO2 < 100, n | 734 |
| Available SpO2 measurements per unique patient, N | 9,302 |
| 1 measurement, n | 8797 |
| 2 measurements, n | 433 |
| 3 measurements, n | 58 |
| 4 measurements, n | 9 |
| 5 measurements, n | 4 |
| 6 measurements, n | 0 |
| 7 measurements, n | 1 |

**Supplementary Table e1. Subject Characteristics in the 7 features models**. The 7 feature models captured 9,990 ICU events from 9,302 unique patients. Variables included in the 7 features machine learning models are SpO2, FiO2, TV, MAP, temperature, PEEP and vasopressor administration. ^*^For subjects older than 89 years, the age was assigned as 90 years of age.

Abbreviations: TV = Title Volume; MAP = Mean arterial pressure; PEEP= Positive end- expiratory pressure
