## Supplementary Table e2 for "Imputation of PaO2 from SpO2 values from the MIMIC-III Critical Care Database Using Machine-Learning Based Algorithms"

|  | **Entire Dataset 1**  **(9,900 events)** | | **Subset 1 (SpO2 < 97%)**  **(1,390 events)** | |
| --- | --- | --- | --- | --- |
|  | **RMSE** | **BIC** | **RMSE** | **BIC** |
| **Neural Network** | **86.2** | **8873.1** | **66.6** | **1196.8** |
| **Linear Regression** | 92.1 | 9002.9 | 67.1 | 1199.0 |
| **Support Vector Regression** | 87.5 | 8902.3 | 68.5 | 1203.5 |
| **Log-linear** | 130.2 | NA | 73.1 | NA |
| **Non-linear** | 97.7 | NA | 82.4 | NA |

**Supplementary Table e2. RMSE and BIC of the 7 features machine learning models compared to published methods**. The RMSE and BIC for the 7 features models were calculated for the entire dataset (9,900 ICU events) and a subset of the dataset with SpO2 < 97% (1,390 ICU events) and compared to the published log-linear and non-linear models.

Abbreviations: RMSE = Root Mean Square Error; BIC = Bayesian Information Criterion; NA = not applicable.
