## Supplementary Table e3 for "Imputation of PaO2 from SpO2 values from the MIMIC-III Critical Care Database Using Machine-Learning Based Algorithms"

|  | **Entire Dataset 1**  **(9,900 events)** | | | | | **Subset 1 (SpO2 < 97%)**  **(1,390 events)** | | | | |
| --- | --- | --- | --- | --- | --- | --- | --- | --- | --- | --- |
|  | **Neural**  **Network** | **Logistic Regression** | **SVM** | **Log-linear** | **Non-linear** | **Neural Network** | **Logistic regression** | **SVM** | **Log-linear** | **Non-linear** |
| **Total, No.** | 9,900 | 9,900 | 9,900 | 9,900 | 9,900 | 1,390 | 1,390 | 1,390 | 1,390 | 1,390 |
| **Sensitivity** | 0.96 | 0.98 | 0.98 | 0.80 | 0.93 | 0.79 | 0.84 | 0.81 | 0.82 | 0.53 |
| **Specificity** | 0.43 | 0.28 | 0.33 | 0.62 | 0.52 | 0.77 | 0.68 | 0.71 | 0.65 | 0.92 |
| **Positive**  **LR** | 1.71 | 1.37 | 1.48 | 2.12 | 1.94 | 3.73 | 2.68 | 2.91 | 2.36 | 6.37 |
| **Negative**  **LR** | 0.09 | 0.07 | 0.06 | 0.33 | 0.13 | 0.28 | 0.24 | 0.27 | 0.27 | 0.51 |
| **Diagnostic**  **OR** | 20.08 | 18.96 | **25.48** | 6.43 | 15.04 | **13.45** | 11.42 | 10.83 | 8.67 | 12.40 |
| **AUROC** | **0.84** | 0.82 | 0.79 | NA | NA | 0.85 | 0.84 | **0.85** | NA | NA |
| **F1** | **0.93** | 0.93 | **0.93** | 0.85 | 0.92 | **0.79** | **0.79** | 0.79 | 0.78 | 0.66 |
| **BIC** | **-2312.40** | -2212.10 | -2243.50 | NA | NA | -221.30 | -216.40 | **-222.30** | NA | NA |

**Supplementary Table e3. Prediction performance of machine learning classification models based on 7 features.** Prediction performance statistics were calculated for the machine learning models based on 7 features and compared to the Log-linear and Non-linear methods for the entire dataset (9,900 ICU events) and for a subset of the events where SpO2 <97% (1,390 ICU events). Variables included in the 7 features machine learning models are SpO2, FiO2, TV, MAP, temperature, PEEP and vasopressor administration.

Abbreviations: SVM = Support Vector Machine; Positive LR = Positive Likelihood Ratio: Negative LR = Negative Likelihood Ratio; Diagnostic OR = Diagnostic Odds Ratio (Ratio of Positive Likelihood Ratio/ Negative Likelihood Ratio); AUROC = Area Under Receiver Operating Characteristic Curve; F1= F1 score; BIC = Bayesian Information Criterion; NA = Not applicable.
