## Supplementary Figure e1 for "Imputation of PaO2 from SpO2 values from the MIMIC-III Critical Care Database Using Machine-Learning Based Algorithms"

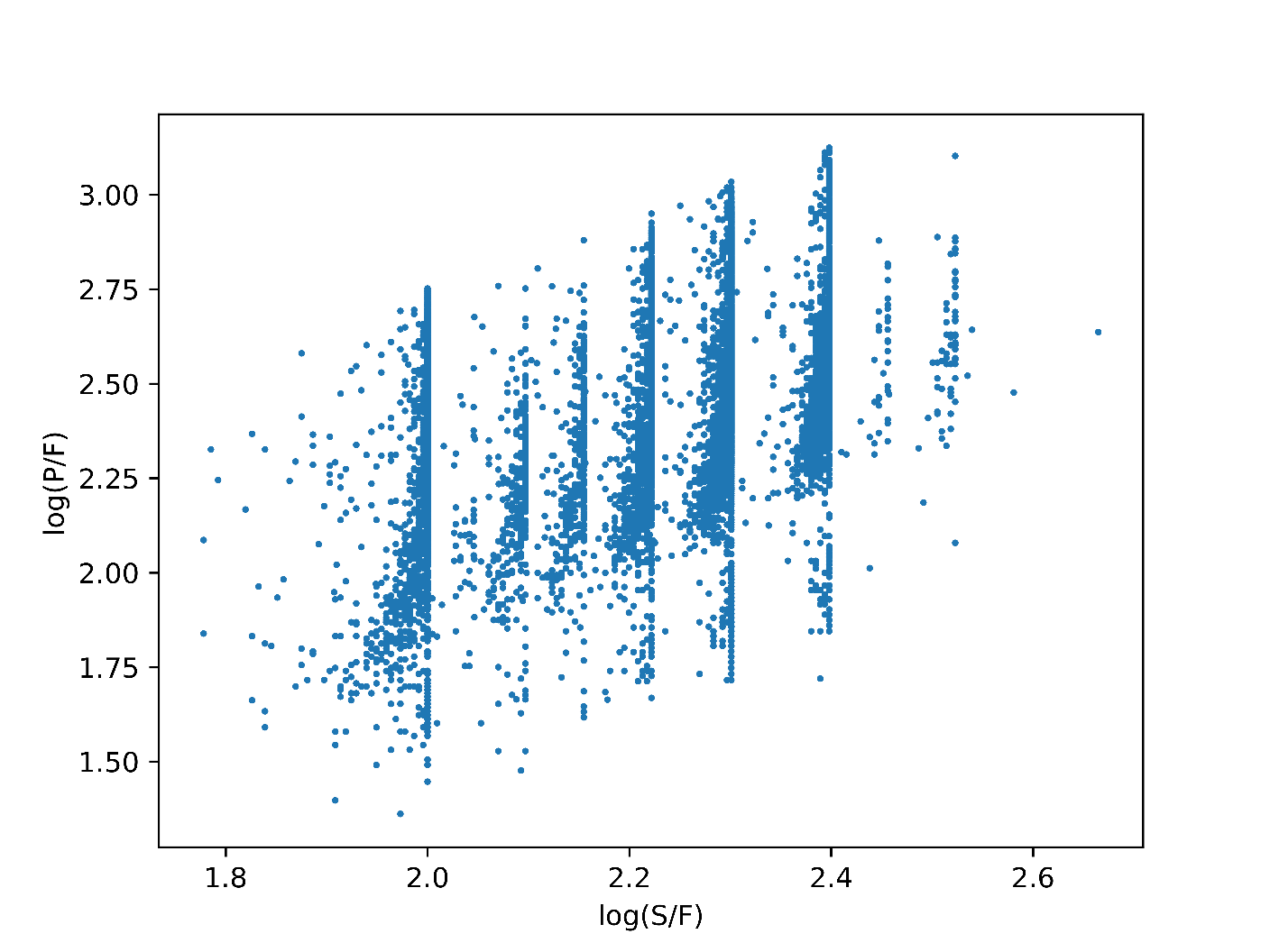

**Supplementary Figure e1. Log linear regression between SpO2/FIO2 (S/F) and PaO2/FIO2 (P/F).** The log-linear relationship between the transformed logarithmic value of the SF and PF ratios is shown. Dataset 1 includes 9,900 ICU events. R^2^ = 0.21. Each point represents a unique ICU event.
