## Supplementary Figure e2 for "Imputation of PaO2 from SpO2 values from the MIMIC-III Critical Care Database Using Machine-Learning Based Algorithms"

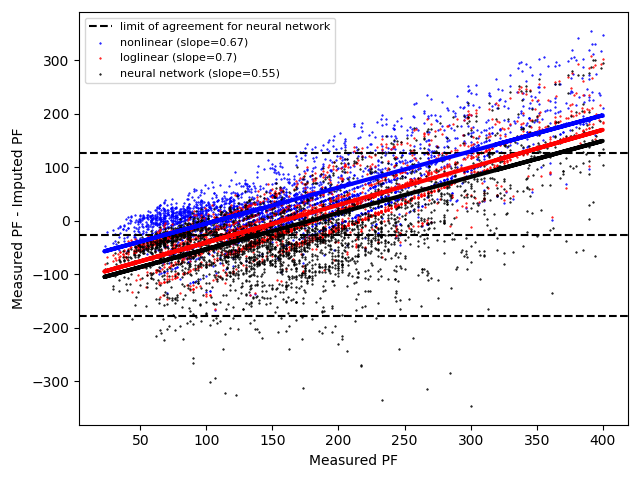

**Supplementary Figure e2: Bland-Altman Plots of imputed versus measured PaO2/FIO2 ratios comparing the published equations and the 3 features Neural Network machine-learning algorithm for Subset 2 (SpO2 < 97%).** Bland-Altman plots of imputed versus measured PaO2/FiO2 ratio comparing the Neural Network machine learning algorithm using 3 features to the published log-linear and non-linear equations. Subset 2 of the entire dataset 2 where SpO2 <97% (3,280 ICU events) is shown and the slope of each least squared line is reported. Each point represents a single ICU event.
